# Physical activity reduces the effect of adiposity genetic liability on hypertension risk in the UK Biobank cohort

**DOI:** 10.1101/2023.09.22.23295992

**Authors:** Chukwueloka Hezekiah, Alexandra I Blakemore, Daniel P Bailey, Raha Pazoki

**Author notes:** **Corresponding author:** Raha Pazoki, Cardiovascular and Metabolic Research Group, College of Health and Life Sciences, Brunel University London, UB8 3PH, United Kingdom ORCID: 0000-0002-5142-2348. **Authors Email Address:** Chukwueloka Hezekiah Dr Raha Pazoki Prof. Alexandra Blakemore Dr Daniel Bailey.

## Abstract

**Background and Purpose:** Hypertension is a leading risk factor for cardiovascular disease (CVD) and is modulated by genetic variants. This study aimed to assess the effect of gene and environmental interaction focusing on adiposity genetic liability and physical activity on hypertension among European and African ancestry individuals within the UK Biobank (UKB).

**Methods:** Participants were 230,115 individuals of European ancestry and 3,239 individuals of African ancestry from UKB. Genetic liability for adiposity were estimated using previously published data including the list of genetic variants and effect sizes for body mass index (BMI), waist-hip ratio (WHR) and waist circumference (WC) using Plink software. The outcome was defined as stage 2 hypertension (systolic blood pressure ≥ 140 mmHg, diastolic blood pressure ≥ 90 mmHg, or the use of anti-hypertensive medications). The association between adiposity genetic liability and the outcome was assessed across categories of self-reported physical activity using logistic regression.

**Results:** Among European ancestry participants, there was up to a 20% hypertension risk difference between participants with a combination of high genetic liability and low physical activity compared with participants with a combination of low genetic liability and high physical activity (*P* <0.001). There was an interaction effect of physical activity on the association between BMI genetic liability and hypertension (*P* _interaction_=0.04). There was no evidence of an association between adiposity genetic liability and hypertension in individuals of African ancestry (*P* > 0.05).

**Conclusion:** This study suggests that engaging in physical activity may reduce the risk of stage 2 hypertension among European ancestry individuals who carry high genetic liability for adiposity. This cannot be inferred for individuals of African ancestry, possibly due to the low African ancestry sample size within the UKB.

## Introduction

Hypertension is a major worldwide health challenge, accounting for an estimated 10.4 million deaths annually ^1^. Approximately 1.39 billion adults suffered from hypertension in 2010 ^2^, with an estimated increase to 1.56 billion people by 2025 ^3^. An estimated 31% of European ancestry adults suffer from hypertension with only 49% hypertension control rate ^4^. The prevalence of hypertension (45%) is higher in African ancestry adults and only 39% hypertension control rate ^4^.

A number of factors are associated with the risk of hypertension including adiposity ^5–7^, lifestyle behaviours such as physical inactivity ^8–11^, and genetics [4]. Adiposity and physical activity are both associated with hypertension. Adiposity increases hypertension ^12,13^, while physical activity, such as aerobic and resistance exercise, effectively reduces hypertension ^8,14^. There is also evidence that self-reported physical activity reduces the effect of adiposity on hypertension risk by 37% in a cohort study of over 13,000 Australian women participants ^15^. Therefore, evidence shows that physical activity can reduce the effect that adiposity has on risk of hypertension.

A genetic susceptibility is recognised for hypertension as numerous genetic factors are linked to it ^16^. For example, adiposity genetic liability increases risk of hypertension ^13^ and improves performance of prediction models of hypertension ^17,18^. A combination of physical activity and low adiposity genetic liability reduces obesity ^19,20^, but it is unclear if physical activity affects the relationship between adiposity genetic liability and hypertension. Understanding the interplay between these different factors (i.e. adiposity, physical activity, and genetic factors) in relation to hypertension risk will further knowledge to improve public health practices in primary prevention of hypertension. The aim of this study was to investigate the effect of adiposity genetic liability on hypertension across different levels of physical activity within European and African ancestry samples of the UK Biobank (UKB).

## Methods

### Ethical Approval

The UKB obtained ethical approval from the North West Multi-centre Research Ethics Committee as a Research Tissue Bank approval. All participants gave informed consent. This research is performed using UKB data under application number 60549. Ethical approval for the current analysis to work on secondary data from the UKB was obtained from Brunel University London, College of Health, Medicine and Life Sciences Research Ethics Committee (reference 27684-LR-Jan/2021-29901-1).

### Study Population

The UKB is a large population-based cohort study established in 2006 to enable comprehensive investigations of genetic, environmental and lifestyle determinants of health, morbidity, and mortality. The study includes >500,000 participants who live in the United Kingdom aged between 40 and 69 years at the time of recruitment ^21^.

### Genotyping and Imputation

DNA extraction and genotyping were undertaken by the UKB. Detailed information regarding genotyping and imputation has been provided elsewhere ^22–24^. In brief, participant blood samples were collected at the UKB assessment centre and genetic data of 488,377 participants was extracted. The first batch of the participants (n=49,950) were genotyped using a comparable Applied Biosystems™ (UK BiLEVE Axiom™ Array by Affymetrix) consisting of 807,411 markers. The remainder of the samples (n=438,427) were genotyped using an Applied Biosystems™ (UKB Axiom™ Array) including 825,927 markers designed to capture short insertions and deletions (indels) and Single Nucleotide Polymorphisms (SNPs) ^22^.

To maximise the use of haplotypes with British and European ancestry for imputation, genotype imputation used three reference panels (Haplotype Reference Consortium, UK10K, and 1000 Genomes phase 3). Genotype imputation was performed by the UKB using the IMPUTE 4 programme. Genetic principal components were computed by the UKB to account for population stratification ^22^.

### Sample for analysis

The current study was performed using two subsets of unrelated individuals of European and African ancestry within the UKB. Participants were excluded (Supplementary Figure 1) if they had withdrawn consent from UKB (n=80). Genetic data were available for 488,377 individuals. After merging genetic and phenotype data, 487,206 individuals remained. Participants who were first- and second-degree relatives (kinship coefficient threshold <0.1768) for at least one other UKB participant (n=26,124) were excluded.Participants were also excluded if (1) they self-reported to be of non-European and non-African ancestry (n=19,743), (2) their self-reported sex did not match their genetic sex (n=330), and (3) they were pregnant or unsure of their pregnancy status at baseline (n=567). In addition, participants who did not declare their smoking status (n=238), or participants with missing data in their *pack-years of smoking* (n=67,115), and current or previous smokers (n=1,065) for whom zero *pack-years of smoking* was calculated were excluded. This could have occurred due to missing values in (1) the age of smoking initiation or cessation, or (2) in the number of cigarettes they smoked per day.

This study further excluded 13,907 individuals who were unsure about their dietary intake of fish, meat, fruits or vegetables and 77,792 individuals with missing data in the main study covariates (see below in Assessment of covariates). In addition, participants who (1) used cholesterol-lowering medication (n=46,679), (2) whose self-reported ancestry did not match their genetic ancestry (n=173), or (3) did not declare drinking status (n=98) were also excluded. The sample was then divided into European (n=230,136) and African (n=3,239) ancestry subsets using self-reported ethnicity data. Participants who withdrew their consent after the analysis was completed were also excluded from the European sample (n=21) and the results were adjusted. The final European sample used in the analysis was 230,115 (Supplementary Figure 1).

### Phenotypic Data

Following informed consent, a broad selection of phenotypic information was collected during baseline assessment in the UKB (the first visit for each participant, which took place between 2006 and 2010). Data were collected from answers provided during interviews and using touch-screen questionnaires. This included socio-demographic, health and lifestyle-related information. Participants also completed a range of physical and anthropometric measurements during the baseline assessment. They provided saliva, urine, and blood samples, which were used for various proteomic, genetic and metabolomic analyses ^25^.

### Blood Pressure and Definition of Hypertension

Two automated systolic blood pressure (SBP) and diastolic blood pressure (DBP) readings were taken after two minutes rest with an Omron HEM 7015-T automated digital blood pressure device. This was done using an appropriately sized cuff on each participant’s left upper arm (the right arm was used where it was not practical to use the left arm). A manual sphygmomanometer was used in instances where the automated blood pressure device could not be used. All blood pressure values were measured in mmHg. Average SBP and DBP were calculated in this study from the recorded readings. For participants with one manual and one automated reading, the average SBP and DBP were calculated using those values only. To minimise the effects of a treatment regime compromising the data for participants on blood pressure-lowering medications, the current study added 15 mmHg to the SBP and 10 mmHg to the DBP ^26^ (n=91,785). In line with the American Heart Association guidelines, hypertension was defined as stage 2 i.e. SBP ≥ 140 or DBP ≥ 90 mmHg^27^. In addition, participants using antihypertensive medication were considered as stage 2 hypertensive.

### Physical Activity Categories

Within the UKB, an adapted version of the short International Physical Activity Questionnaire (IPAQ) was used to assess physical activity using a touch screen self-reported questionnaire ^28^. Information on the frequency, intensity and duration of walking, moderate and vigorous physical activity was collected. Cassidy and colleagues ^29^ used the IPAQ data processing guidelines to produce the Metabolic Equivalent of Task (MET) minutes per week of physical activity by multiplying the duration of walking, moderate and vigorous physical activities by 3.3, 4.0 and 8.0 METs, respectively. These were summed to provide total physical activity in MET min/week ^30^. The data was returned to the UKB by Cassidy and colleagues ^29^, which were used in the present study to categorise physical activity into (1) low (total physical activity < 600 MET-minutes/week) and (2) moderate (≥ 600 MET-minutes/week) or high (≥ 3,000 MET-minutes/week) physical activity ^30^. We grouped moderate and high physical activity together as the World Health Organization’s recommends performig moderate or high physical activity to gain health benefits ^31^.

### Assessment of covariates

Dietary intake was assessed using a self-report questionnaire. Participants were asked to select their daily quantity of dietary consumption using Assessment Centre Environment touchscreen multiple-choice questions. This included *vegetable* (*cooked* and *raw*), *fruit* (*fresh* and *dried*), *oily fish* and *meat* (*processed* and *unprocessed*, including *poultry*, *lamb*, and *pork*) intake.

*Smoking status* in the UKB was assessed using a self-reported question categorising smoking status into current, past, and never smoking. *Pack-years of smoking* was available for European (n=230,115) and African (n=3,239) ancestry samples. *Pack-years of smoking* was assessed as the number of cigarettes smoked per day divided by twenty (as the average pack size), multiplied by the number of years smoking (https://biobank.ndph.ox.ac.uk/showcase/field.cgi?id=20161; access date 29 June 2023). The number of years smoking was calculated by subtracting the age in which the participant started smoking from the age they stopped smoking. In this current study, smoking was defined using the combination of both *Smoking status* and *Pack-years of smoking.* Current or previous smokers with any values in their *Pack-years of smoking* were categorised as smokers. Never smokers were considered as participants who reported never smoked and had zero values in *Pack-years of smoking*.

Additional covariates for the analysis included self-reported alcohol drinking status (current, past and non-drinkers) and low-density lipoprotein (LDL) cholesterol measured by enzymatic protective selection analysis on a Beckman Coulter AU5800. Further details of quality control and sample preparation for the UKB biomarker data have been published previously ^32^.

### Adiposity genetic liability

In the current study, adiposity genetic liability was generated using previously reported genetic variants for European and African ancestry individuals. These variants were associated with body mass index (BMI) ^33,34^, waist-hip ratio (WHR) ^35,36^, or waist circumference (WC) ^35,36^. Within the European ancestry sample, a list of 155 SNPs with their weights (β-coefficients) from a GWAS performed by Winkler and colleagues ^33^ was used to estimate the BMI genetic liability. β-coefficients for WHR SNPs (n=27) and WC SNPs (n=41) were obtained from Shungin and colleagues ^35^ and were used to estimate WHR and WC genetic liabilities. To estimate BMI genetic liability for individuals of African ancestry, a list of seven SNPs with β-coefficients from Ng and colleagues ^34^ was used. To estimate WHR and WC genetic liability in African ancestry, a list of three WHR and two WC SNPs with β-coefficients from Liu and colleagues ^36^ were used.

As part of the SNP selection process in this study, SNPs that (1) did not reach a GWAS significance threshold of P < 5 x 10^-8^, (2) had Minor allele frequency <0.01, (3) were dependent to other SNPs as demonstrated by linkage disequilibrium (LD) parameter (*R*^2^>0.1) were excluded. The final list of SNPs used in the analyses are presented in Supplementary Tables 1 and 2.

Using Plink v1.9 ^37^ to calculate genetic liabilities, the previously estimated effect of each genetic variant on an adiposity phenotype (i.e. BMI, WHR, and WC) were multiplied by the number of risk alleles each UKB participant carried on the BMI, WHR and WC SNPs. The products were then summed across all SNPs to produce overall weighted genetic liability for BMI, WHR and WC for each participant. The weighted genetic liabilities for each participant were standardised by subtracting the average genetic liability within the sample from the participant’s genetic liability, divided by standard deviation of the genetic liability within the sample.

### Data Presentation and Statistical Analysis

To identify genetic distance of the participants, a cluster analysis was conducted using the K-means algorithm ^38^. The K-means algorithm requires a parameter specifying the number of clusters (K) ^39^. A ‘K’ value of seven was assigned according to the number of categories within the UKB self-reported phenotypic variable, *ethnic background* (Supplementary Table 3). The genetically derived clusters were compared with the self-reported ancestries to identify participants whose self-reported ancestry did not match with the genetically derived ancestry.

Logistic regression was used to estimate the risk of stage 2 hypertension per unit increase in the standardised adiposity genetic liability (BMI, WHR and WC). This was estimated for the whole sample and within separate physical activity categories (low and moderate/high). The combined effect of adiposity genetic liability and physical activity was examined by comparing the risk of stage 2 hypertension in each combined category with the reference group (low genetic liability and moderate/high physical activity).

The analyses were adjusted for age, sex, daily alcohol intake, smoking status, daily fruit and vegetable intake, meat intake, fish intake, and LDL cholesterol. For genetic liabilities that demonstrated a statistically significant association with stage 2 hypertension within the whole sample, an interaction test was performed to identify if physical activity could modify the effect of genetic liability on stage 2 hypertension. Statistical significance was accepted if the type 1 error (*P*-value) was below 0.05. All statistical analyses were implemented in R v4.0.0 ^40^.

## Results

### European ancestry results

This study included 230,115 European ancestry participants of the UKB (Table 1). Prevalence of hypertension was significantly different between men (52.90%) and women (40.30%; *P* < 0.001). No differences were observed between men and women in terms of MET physical activity.

**Table 1.** Baseline characteristics of the study sample.

| Characteristics | African |  |  | European |  |  |
| --- | --- | --- | --- | --- | --- | --- |
|  | Female<br>N=1959 | Male<br>N=1280 | P-value for sex<br>difference<sup>†</sup> | Female<br>N=128,277 | Male<br>N=101,838 | P-value for<br>sex<br>difference<sup>†</sup> |
| AGE (Years) |  |  |  |  |  |  |
| Mean (SD) | 50.90 (7.34) | 50.30 (7.68) | 0.04 | 55.40 (7.90) | 55.40 (8.13) | 0.04 |
| Median [Min, Max] | 50 [40, 70] | 49 [40, 70] |  | 56 [40, 70] | 56 [38, 73] |  |
| Hypertension cases by AHA guidelines * |  |  |  |  |  |  |
| NO, n (%) | 962 (49.10%) | 617 (48.20%) | 0.64 | 76,612 (59.70%) | 47,963 (47.10%) | <0.001 |
| YES, n (%) | 997 (50.90%) | 663 (51.80%) |  | 51,665 (40.30%) | 53,875 (52.90%) |  |
| Levels of Physical Activity |  |  |  |  |  |  |
| Low, n (%) | 419 (21.40%) | 281 (22.00%) | 0.74 | 23,099 (18.00%) | 18,505 (18.20%) | 0.31 |
| Moderate/High, n (%) | 1540 (78.60%) | 999 (78.00%) |  | 105,178 (82.00%) | 83,333 (81.80%) |  |
| LDL Cholesterol (mmol/L) |  |  |  |  |  |  |
| Mean (SD) | 3.32 (0.78) | 3.38 (0.83) | 0.07 | 3.72 (0.84) | 3.70 (0.78) | <0.001 |
| Median [Min, Max] | 3.27 [0.85, 7.08] | 3.32 [1.29, 7.06] |  | 3.67 [0.80, 9.74] | 3.67 [0.27, 8.99] |  |
| Smoking Status |  |  |  |  |  |  |
| Non-Smoker, n (%) | 1650 (84.20%) | 970 (75.80%) | <0.001 | 88,922 (69.30%) | 63,638 (62.50%) | <0.001 |
| Smoker, n (%) | 309 (15.80%) | 310 (24.20%) |  | 39,355 (30.70%) | 38,200 (37.50%) |  |
| Systolic Blood Pressure (mmHg) |  |  |  |  |  |  |
| Mean (SD) | 139 (21.80) | 142 (19.20) | <0.001 | 136 (20.50) | 142 (18.60) | <0.001 |
| Median [Min, Max] | 136 [91, 255] | 139 [103, 236] |  | 133 [72, 253] | 140 [85, 268] |  |

Table 1. Baseline characteristics of the study sample.
|  |  |  |  |  |  |  |
| --- | --- | --- | --- | --- | --- | --- |
| Diastolic Blood Pressure (mmHg)<br>Mean (SD)<br>Median [Min, Max] | 86.90 (12.30)<br>86 [53.50, 134] | 86.70 (12)<br>86 [55, 129] | 0.63 | 81.60 (10.90)<br>80.50 [45, 142] | 85.60 (10.90)<br>85 [42, 148] | <0.001 |
| Takes blood pressure lowering medication<br>NO, n (%)<br>YES, n (%) | 1466 (74.80%)<br>493 (25.20%) | 1030 (80.50%)<br>250 (19.50%) | <0.001 | 114,369 (89.20%)<br>13,908 (10.80%) | 89,337 (87.70%)<br>12,501 (12.30%) | <0.001 |
| Alcohol Status<br>Never, n (%)<br>Previous, n (%)<br>Current, n (%) | 355 (18.10%)<br>80 (4.10%)<br>1524 (77.80%) | 176 (13.80%)<br>76 (5.90%)<br>1028 (80.30%) | <0.001 | 5272 (4.10%)<br>4161 (3.20%)<br>118,844 (92.60%) | 1811 (1.80%)<br>3002 (2.90%)<br>97,025 (95.30%) | <0.001 |
| Daily Fruit and Vegetable intake<br>Mean (SD)<br>Median [Min, Max] | 9.15 (5.78)<br>8 [0, 53] | 8.60 (7.40)<br>7 [0, 72] | 0.03 | 8.39 (4.39)<br>8 [0, 106] | 7.28 (4.54)<br>6.50 [0, 13] | <0.001 |
| Oily Fish Intake<br>Mean (SD)<br>Median [Min, Max] | 1.96 (0.99)<br>2 [0, 5] | 1.89 (1.01)<br>2 [0, 5] | 0.05 | 1.66 (0.91)<br>2 [0, 5] | 1.58 (0.92)<br>2 [0, 5] | <0.001 |
| Meat Intake<br>Mean (SD)<br>Median [Min, Max] | 7.41 (3.04)<br>7 [0, 23] | 8.90 (3.20)<br>9 [0, 25] | <0.001 | 7.34 (2.81)<br>8 [0, 25] | 8.43 (2.66)<br>9 [0, 25] | <0.001 |
| BMI (kg/m<sup>2</sup>)<br>Mean (SD)<br>Median [Min, Max] | 29.90 (5.71)<br>29.20 [18.20, 68.10] | 28.30 (4.18)<br>27.90 [17.70, 57.5] | <0.001 | 26.50 (4.88)<br>25.60 [12.10, 66.20] | 27.30 (4.03)<br>26.90 [12.80, 61.70] | <0.001 |

Statistical analysis were performed using the chi-squared test, for categorical
variables and student t-test, for numerical variables. \* Systolic blood pressure >140
or diastolic blood pressure >90. † P-value for sex difference was obtained using
logistic regression adjusted for age, sex, physical activity, smoking status, alcohol
status, meat and fish intake, fruit and vegetable intake, and Low-density lipoprotein
cholesterol. AHA, American Heart Association; SD, standard deviation; min,
minimum; max, maximum; LDL, Low-density lipoprotein.

Table 2 shows the effect of adiposity genetic liability, as a continuous variable, on the risk of stage 2 hypertension across categories of physical activity. Each unit increase in the standardised adiposity genetic liabilities (BMI, WHR, and WC) were significantly associated with hypertension (OR_BMI_=1.05; OR_WHR_=1.04; OR_WC_=1.04) within the whole sample (Table 2), as well as within the low (OR_BMI_ = 1.08; OR_WHR_=1.05; OR_WC_= 1.06) and moderate/high physical activity subgroups (OR_BMI_ = 1.05; OR_WHR_= 1.03; OR_WC_= 1.04). A statistically significant interaction was observed for physical activity reducing the association between BMI genetic liability and hypertension (*P* _interaction_=0.04; Table 2). This interaction was not statistically significant for the other adiposity genetic liabilities (WHR or WC).

**Table 2.**
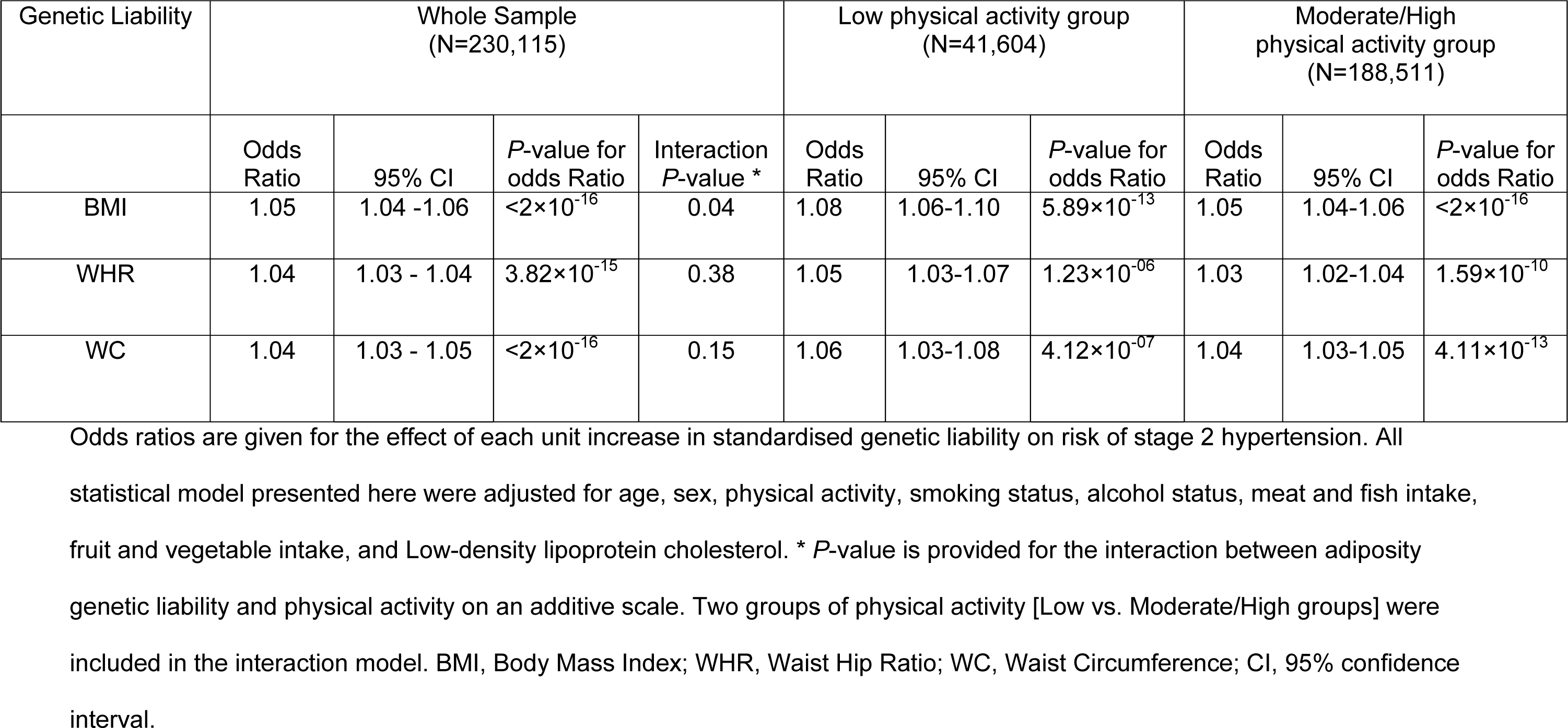
Association between hypertension and adiposity genetic liability in European Ancestry sample.

| Genetic Liability | Whole Sample<br>(N=230,115) |  |  |  | Low physical activity group<br>(N=41,604) |  |  | Moderate/High<br>physical activity group<br>(N=188,511) |  |  |
| --- | --- | --- | --- | --- | --- | --- | --- | --- | --- | --- |
|  | Odds<br>Ratio | 95% CI | *P*-value for<br>odds Ratio | Interaction<br>*P*-value * | Odds<br>Ratio | 95% CI | *P*-value for<br>odds Ratio | Odds<br>Ratio | 95% CI | *P*-value for<br>odds Ratio |
| BMI | 1.05 | 1.04 -1.06 | $<2 \times 10^{-16}$ | 0.04 | 1.08 | 1.06-1.10 | $5.89 \times 10^{-13}$ | 1.05 | 1.04-1.06 | $<2 \times 10^{-16}$ |
| WHR | 1.04 | 1.03 - 1.04 | $3.82 \times 10^{-15}$ | 0.38 | 1.05 | 1.03-1.07 | $1.23 \times 10^{-06}$ | 1.03 | 1.02-1.04 | $1.59 \times 10^{-10}$ |
| WC | 1.04 | 1.03 - 1.05 | $<2 \times 10^{-16}$ | 0.15 | 1.06 | 1.03-1.08 | $4.12 \times 10^{-07}$ | 1.04 | 1.03-1.05 | $4.11 \times 10^{-13}$ |

Odds ratios are given for the effect of each unit increase in standardised genetic liability on risk of stage 2 hypertension. All statistical model presented here were adjusted for age, sex, physical activity, smoking status, alcohol status, meat and fish intake, fruit and vegetable intake, and Low-density lipoprotein cholesterol. \* *P*-value is provided for the interaction between adiposity genetic liability and physical activity on an additive scale. Two groups of physical activity [Low vs. Moderate/High groups] were included in the interaction model. BMI, Body Mass Index; WHR, Waist Hip Ratio; WC, Waist Circumference; CI, 95% confidence interval.

Compared with participants with a combination of low genetic liability and moderate/high physical activity (Table 3), the largest risk of stage 2 hypertension was observed among participants with a combination of low physical activity and high adiposity genetic liability (OR_BMI_=1.20; OR_WHR_=1.15; OR_WC_=1.17).

**Table 3.**
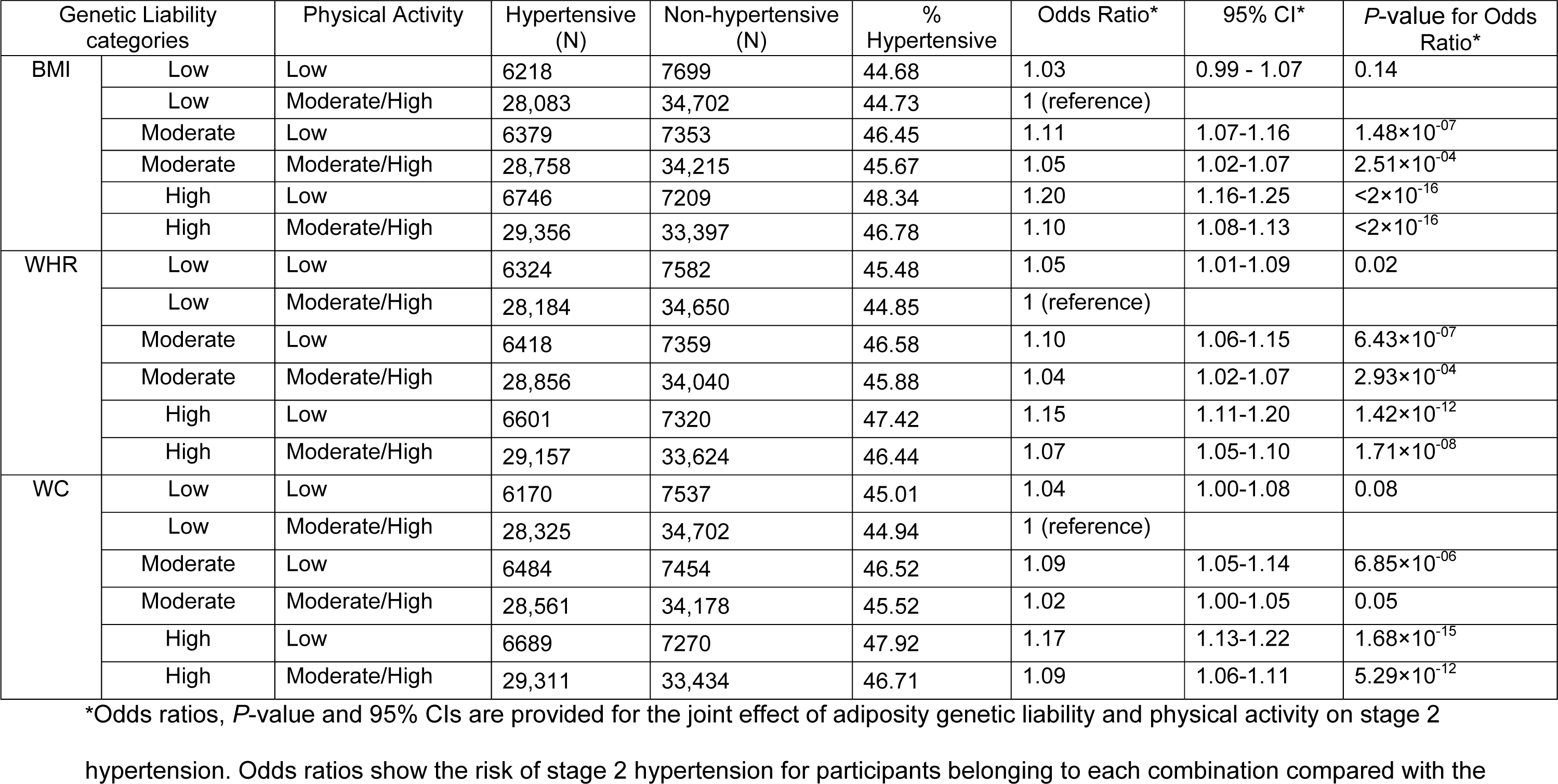
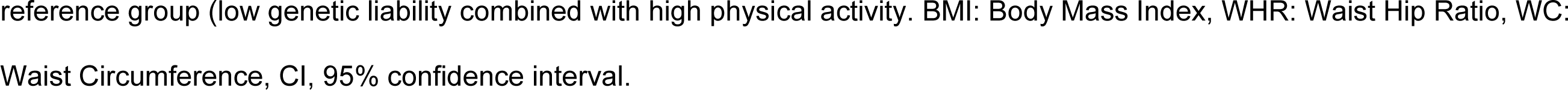
Prevalence of stage 2 hypertension across different physical activity and adiposity genetic liability groups in individuals of European ancestry in the UKB.

### African ancestry results

Within the African ancestry sample, 3239 participants were included (Table 1). There were no significant difference between men and women in terms of (1) prevalence of hypertension (*P* = 0.64) and (2) physical activity (*P* = 0.74). None of the adiposity genetic liabilities was significantly associated with hypertension within the whole sample or within any of the physical activity level groups (Table 4).

**Table 4.** Association between hypertension and adiposity genetic liability in African Ancestry.

| Genetic Liability | Whole Sample<br>(N=3239) |  |  |  | Low physical activity group<br>(N=700) |  |  | Moderate or High physical<br>activity group<br>(N=2539) |  |  |
| --- | --- | --- | --- | --- | --- | --- | --- | --- | --- | --- |
|  | Odds<br>Ratio | 95% CI | *P*-value for<br>odds Ratio | Interaction<br>*P* value | Odds<br>Ratio | 95% CI | *P*-value for<br>odds Ratio | Odds<br>Ratio | 95% CI | *P*-value for<br>odds Ratio |
| BMI | 1.06 | 0.98-1.15 | 0.16 | NA | 1.06 | 0.89-1.27 | 0.50 | 1.06 | 0.97-1.16 | 0.22 |
| WHR | 1.01 | 0.94-1.09 | 0.80 | NA | 0.94 | 0.81-1.10 | 0.46 | 1.03 | 0.95-1.12 | 0.46 |
| WC | 0.97 | 0.88-1.06 | 0.49 | NA | 0.92 | 0.75-1.12 | 0.39 | 0.98 | 0.88-1.09 | 0.69 |

Odds ratios are given for the effect of each unit increase in standardised genetic liability on risk of stage 2 hypertension within the African sample. Model was adjusted for age, sex, physical activity, smoking status, alcohol status, meat and fish intake, fruit and vegetable intake, and Low-density lipoprotein cholesterol. BMI, Body Mass Index; WHR, Waist Hip Ratio; WC, Waist Circumference; CI, 95% confidence interval.

## Discussion

The main findings of this study are that (1) adiposity genetic liabilities increase risk of hypertension, and (2) physical activity could offset the risk of hypertension caused by BMI genetic liability in European ancestry.

The combined effect of obesity and physical activity on the risk of hypertension has been investigated in non-genetic based studies. Jackson and colleagues ^15^ reported that the risk of hypertension was 37% higher in healthy weight inactive Australian women compared with healthy weight highly active women. In a Chinese population, participants with obesity and low levels of self-reported physical activity had a higher risk of hypertension compared with participants with normal weight and high physical activity levels ^41^. The present study was focused on adiposity genetic liabilities rather than phenotypes, which has the potential to identify risk of hypertension early in the life course and the potential importance of engaging in physical activity. The findings here imply that interventions targeting physical activity could be beneficial in reducing the risk of hypertension in individuals who are genetically predisposed to adiposity. Among individuals who are not genetically predisposed to adiposity, physical activity does not seem to play a role in changing the risk of hypertension.

In the current study, different combinations of levels of adiposity genetic liability and physical activity were compared with the optimal combination being low genetic liability and moderate/high physical activity. We found that in European ancestry participants all combinations increased risk of hypertension (up to 20% risk difference) compared with the optimal. The effect of these combinations on hypertension were smaller in the current study compared with the effects observed in the studies by Jackson and colleagues ^15^ and Li and colleagues ^41^ that focused on obesity phenotype and did not include genetics. They also were focused on specific sub populations (women or obese subjects). The use of genetics in our study allows risk estimation across the lifecourse. In addition our study was performed in a sample of generally healthy men and women which provides a more generalisable results to the healthy population in producing public health programs.

The findings from this present study could inform public health policy in the context of physical activity interventions potentially producing varied effects on hypertension depending on the genetic predisposition of the target population. The present study showed that physical activity could have greater effects in individuals with increased adiposity genetic risk. Further research should investigate the economic and hypertension-related benefits of targeting genetically high-risk groups rather than the whole population in public health interventions.

Genetic predisposition to adiposity was not associated with hypertension among African ancestry participants. Genetic studies in African ancestry populations have revealed no association between adiposity genotype and hypertension ^42^. Shi and colleagues ^42^ used European specific BMI genetic liability to investigate its association with hypertension in a small African sample (n=369) and found no relationship. The results are unexpected as there is evidence that both adiposity ^43^ and low physical activity ^44^ are associated with risk of hypertension in African ancestry individuals. There are differences in fat distribution in African ancestry individuals compared with European ancestry individuals. For example, the body fat mass is lower and the lean muscle mass is higher within the African ancestry individuals as opposed to the European ancestry individuals when compared at the same BMI level ^45^. It might be useful for future studies to consider taking variation in muscle and fat mass distribution into account when analysing data from African ancestry individuals. The fact that the findings from the current study did not identify an association between African ancestry-specific genetic liability and hypertension could also be due to an insufficient sample size. The African ancestry subset of the UKB has a relatively small sample size (n=3,239) compared with the European sample and this could justify why an effect was observed within the European sample and not the African sample. In addition, the low number of SNPs identified within the African ancestry data could also explain the lack of association in the African ancestry sample. Future studies with larger sample sizes are needed to assess the combined effects of adiposity genetic liability and physical activity on hypertension risk among African ancestry individuals.

The present study has several strengths, including the large European ancestry sample size providing statistical power for various physical activity subgroup analyses. The wide array of relevant covariates collected by the UKB enabled adjustments for potential confounding factors, thereby facilitating a more accurate estimation of the risks involved. The current study investigated the combined effect of adiposity genetic liability and physical activity on risk of hypertension. This original approach offers insight to be used in creating preventive strategies, particularly targetting individuals with a high genetic predisposition for adiposity.

A potential limitation of the study is that African populations are both diverse ^46^ and their genetic data relatively understudied in terms of genomics research, so the ability to accurately assess genetic liability may be limited compared with European ancestry populations. Furthermore, the use of genetic liability provides a life course insight and typically explains small variation in outcomes ^47^. Another limitation is that physical activity was assessed via self-reported which has the potential to overestimate or underestimate the true physical activity levels ^48^. Further studies using more objective and precise measure of physical activity such as data obtained using accelerometer could produce a more valid measure of physical activity ^49^.

## Conclusion

The findings of this study demonstrate that increasing adiposity genetic liability is associated with hypertension. Individuals of European ancestry with increased adiposity genetic liability may benefit more from higher levels of physical activity in relation to reduced risk of stage 2 hypertension compared to individuals with lower adiposity genetic liability. The current study highlights that physical activity interventions might help lower the risk of stage 2 hypertension in European ancestry individuals who have a genetic tendency to become overweight and obese. These effects could not be replicated within the African ancestry sample of the UKB. Further research with larger samples and consideration of the muscle and fat mass distribution in the African ancestry is proposed.

## Data Availability

Not applicable

## Acknowledgments

This study has been performed using the UKB application 60549.

## Author Contributions

Conceptualization, R.P.; Data curation, C.H.; Formal analysis, C.H.; Funding acquisition, R.P.; Investigation, C.H.; Methodology, C.H., R.P.; Project administration, R.P.; Resources, R.P.; Supervision, R.P., A.I.B. and D.P.B; Writing— original draft, C.H.; Writing—review & editing, C.H., A.I.B., D.P.B. and R.P. All authors have read and agreed to the published version of the manuscript.

## Institutional Review Board Statement

The study was conducted in accordance with the Declaration of Helsinki and approved by the Institutional Review Board (or Ethics Committee) of Brunel University London, College of Health, Medicine and Life Sciences (27684-LR-Jan/2021-29901-1).

## Informed Consent Statement

Informed consent was obtained from all subjects involved in the study.

## Data Availability Statement

Not applicable.

## Supplemental Material

Supplementary Figure 1

Supplementary Table 1– Supplementary Table 3

References 1–4

## Non-standard Abbreviations and Acronyms

UKB: UK Biobank
SNPs: single nucleotide polymorphisms
SBP: systolic blood pressure
DBP: diastolic blood pressure
IPAQ: international physical activity questionnaire
MET: metabolic equivalent of task

